# Thriving in Place: Multidimensional Neighborhood Typologies and Cognitive Function Among Older Americans

**DOI:** 10.1101/2025.11.01.25339304

**Authors:** Jiao Yu, Weidi Qin, Jiuzhou Wang, Eva Kahana

**Affiliations:** School of Public Health, Yale University, New Haven, CT, USA; Sandra Rosenbaum School of Social Work, University of Wisconsin–Madison, Madison, WI, USA; School of Public Health, University of Minnesota Twin Cities, Minneapolis, MN, USA; Department of Sociology, Case Western Reserve University, Cleveland, OH, USA

## Abstract

**Background:** Neighborhood physical, social, and service environments are increasingly recognized as important contextual factors related to cognitive health; however, few studies have examined how these features collectively shape cognitive outcomes. This study aimed to classify neighborhood typologies based on a constellation of neighborhood features and investigate their associations with cognitive function among US older adults.

**Methods:** We conducted a cross-sectional analysis of 6,480 participants from the 2016 wave of the Health and Retirement Study. Neighborhood features were derived from the 2015 National Neighborhood Data Archive, including measures of neighborhood deprivation, service facilities, food access, healthcare, and environmental hazards. The Partitioning Around Medoids (PAM) machine learning clustering algorithm was used to classify neighborhood typologies. Multilevel regression analyses were performed to examine the associations between neighborhood typologies and cognitive function.

**Results:** Four neighborhood typologies were identified: (1) low deprivation, green neighborhood, (2) mid-SES, high hazard neighborhood, (3) high-amenity neighborhood, and (4) disadvantaged neighborhood. Regression results revealed significant disparities in cognitive function across neighborhood typologies. Older residents in high-amenity neighborhoods characterized by extensive facilities and cognitively stimulating services demonstrated better cognitive function (*β =* 3.85*, 95% CI:* 1.23-6.47) compared to those in disadvantaged, resource-scarce neighborhoods, after adjustment for individual-level characteristics.

**Conclusions:** The identified neighborhood typologies reveal an unequal distribution of amenities and hazards, which may help account for considerable inequities in late-life cognitive health. Tailored community initiatives addressing amenity availability and environmental hazards could be pivotal in promoting cognitive health and supporting aging in place.

## Introduction

Cognitive decline represents a major health challenge in later life. In the United States, mild cognitive impairment affects approximately 22% of adults aged 65 years or older, and approximately 10% of this population lives with dementia [1]. There has been growing recognition that cognitive health disparities should be understood within specific social and environmental contexts, with neighborhood environments playing a particularly important role in shaping cognitive outcomes in later life. Compared to younger adults, older adults spend more time in their residential neighborhoods, and thus the importance of neighborhood environments increases with age [2]. Neighborhoods are primary settings for social interaction, physical activity, and access to resources, thereby potentially facilitating or impeding health-promoting behaviors that can delay or accelerate cognitive aging. Extensive research has pointed to the connection between neighborhood characteristics and cognitive health outcomes [3–6]. However, much of this scholarship has examined isolated environmental features in relation to cognition, such as exposure to green space, food access, or neighborhood walkability [7–9]. While these findings offer valuable insights, few studies have examined how the totality of neighborhood contexts might influence cognitive health. That is, we know little about how the combination of different neighborhood features can collectively influence cognitive function. Understanding how these multidimensional and potentially interactive neighborhood features contribute to health is particularly important, as older adults’ experiences of place often reflect a dynamic interplay of physical, social, and structural conditions.

Neighborhood environments, as social determinants of health, play a significant role in shaping older adults’ cognitive health [4,5]. Empirical evidence has shown that socioeconomic deprivation, infrastructure, and amenities within one’s neighborhood can substantially influence older adults’ cognitive health. For example, significant cognitive decline is associated with living in a neighborhood with a high concentration of socioeconomically disadvantaged households [10,11], a lack of neighborhood walkability [9], and high levels of physical disorder [12]. Additionally, earlier evidence suggests that neighborhoods rich in diverse amenities (i.e., convenience stores, pharmacies, gyms, and recreation centers) may promote community-dwelling older adults’ mobility and participation in social activities [13,14], which in turn can delay cognitive decline and reduce the risk of cognitive impairment [15].

Neighborhood features, which serve as upstream factors, may either delay or accelerate cognitive impairment by shaping daily experiences and exposures related to exercise, diet, pollutants, and social engagement or isolation [16]. Amenity-rich neighborhoods featuring parks, recreation centers, or civic organizations may promote cognitive health by facilitating physical activities, social engagement, and mental stimulation, which are known to build cognitive reserve[4]. Conversely, denser urban neighborhoods with environmental stressors such as air pollutants, noise, crowding, and physical disorder can elicit various levels of chronic stress, which has been shown to have deleterious effects on brain structures critical for memory and learning [17,18]. The existing literature on the neighborhood effect has not yet provided the most plausible mechanisms or the precise directionality of associations between specific neighborhood features and cognitive outcomes. It is likely that the mechanism varies depending on the particular feature under investigation. Moreover, rather than a single explanatory pathway, it is reasonable to assume that multiple mechanisms may operate interactively and reinforce one another to shape cognitive health outcomes [16].

Existing studies have largely examined neighborhood characteristics in isolation, focusing on individual dimensions such as socioeconomic disadvantage, amenities, polluting hazards, etc. [19–21]. While valuable, they fail to elucidate how the combination of different neighborhood features (i.e., both protective and risk factors) may collectively affect cognitive outcomes. Specifically, a neighborhood may contain a mixture of both protective factors that promote cognitive health (e.g., green space, libraries) and risk factors that accelerate cognitive decline (e.g., pollution, high density of highways) [22]. For example, while a high level of concentrated disadvantage in a neighborhood may increase the risk of impaired cognitive function, it is also possible that older adults in socioeconomically disadvantaged neighborhoods experience stronger social ties with neighbors, thus protecting against health deterioration [23].

Additionally, neighborhoods constitute a critical milieu involving participants, physical space, activities, and interactions that collectively imbue a place with meaning [24]. These environments shape opportunities for engagement, access to resources, and exposure to risks, all of which accumulate in synergistic ways to influence well-being in later life [25]. Building on this perspective, the present study draws upon the ecological framework of aging to understand the complex interplay between individuals and their environments in shaping cognitive health [4,24,26]. The ecological theory of aging suggests that successful aging in place is achieved through optimized person-in-environment fit [26]. That is, the balance between personal competence and environmental press plays an important role in late-life well-being. While risk factors within the environment may disrupt this balance by imposing excessive demands, protective factors may enhance person-environment fit by buffering against environmental stressors or by providing resources that support older adults’ capacities. Informed by this framework, we conceptualize neighborhood environments as upstream determinants of health that can constitute both opportunities and barriers to cognitive health. By examining neighborhood-cognition associations with the consideration of multidimensional neighborhood risk and protective factors, our findings will provide an essential basis for developing tailored community interventions aimed at promoting cognitive resilience and supporting healthy aging in place.

Taken together, while prior research has established that individual neighborhood characteristics, such as socioeconomic disadvantage, environmental hazards, or neighborhood walkability, are associated with cognitive health, we know little about how the clustering of both risk and protective factors across multiple neighborhood dimensions collectively influences cognitive outcomes. Recent scholarship has advanced the framework of the exposome, which refers to the totality of environmental exposures that individuals encounter and their influences on health outcomes [27]. This holistic framework motivates our creation of multidimensional neighborhood typologies that provide a more comprehensive and realistic account of concurrent exposures within neighborhood contexts. Building on this premise, the present study aims to investigate multidimensional features of neighborhood environments in relation to cognitive function. Using data from the nationally representative Health and Retirement Study (HRS) and applying a novel machine-learning clustering method, this research has two primary aims. First, we will identify neighborhood typologies based on the dimensions of neighborhood socioeconomic status, environmental hazards, and access to amenities and facilities. Second, we will examine how these empirically derived neighborhood typologies are associated with cognitive function among community-dwelling older adults.

## Methods

### Data

Data were drawn from the Health and Retirement Study (HRS) and the National Neighborhood Data Archive. HRS is a nationally representative longitudinal study of Americans aged 51 and older. Data are collected biannually since 1992, and the sample is replenished every six years. Using a multistage probability sampling design, the HRS collected demographic, socioeconomic, and health data as well as biological subsamples for genomic and biomarker analyses [28]. The present study used data from the participants who completed the 2016 wave core survey and provided salivary DNA samples with available genetic data collected between 2006 and 2012 [29].

Neighborhood data were obtained from the National Neighborhood Data Archive (NaNDA). NaNDA is a publicly available data repository containing nationwide contextual measures of the physical and social environments, e.g., walkability, amenities, green space, and healthcare [30]. The HRS team geocoded respondents’ residential addresses to 2010 US census tract boundaries, allowing us to link the HRS sample to tract-level contextual measures from the NaNDA. We linked the 2016 HRS sample to the 2015 wave of NaNDA data at the census tract level to ensure appropriate temporal ordering; this approach captured neighborhood features one year before the collection of cognitive function measurements. There were 3,352 unique tracts in this study with an average of 2 persons per tract (range: 1-41).

A total of 20,880 participants completed the 2016 wave HRS interview. We restricted our sample to community-dwelling adults aged 65 and older (n=10,104). We excluded 780 participants with incomplete cognition information, 994 participants lacking genomic data, and 827 participants who moved between census tracts during 2015 and 2016. We further excluded 676 participants missing census tract information who could not be matched to the neighborhood data. Among the remaining participants, listwise deletion was applied to those with missing values in key covariates (n = 347). This resulted in a final analytic sample consisting of 6,480 community-dwelling older adults. The sample selection process is shown in Appendix Figure S1.

### Measures

#### Cognitive Function

Cognitive function was assessed using an adapted version of the Telephone Interview for Cognitive Status (TICS), a cognitive scale with a maximum of 27 points [31]. Participants were asked a series of questions assessing (1) immediate and delayed word recall measuring memory (0-20 points); (2) a serial sevens subtraction test measuring working memory (0-5 points); and (3) a counting backwards test measuring mental processing speed (0-2 points). For respondents who were unable to participate, cognitive function was assessed through a proxy interview. In the present study, we excluded proxy respondents and only included cognitive scores from self-respondents. Cognitive function was measured as the sum of the component scores, with higher scores representing better cognitive function.

#### Neighborhood Features

Neighborhood measures came from the NaNDA (2015). Consistent with prior work and data availability, we used census tracts as proxies for neighborhoods [32]. Detailed methodology for the construction of these measures, including item classifications and buffer calculations is available from the NaNDA repository [33]. In this study, we included a comprehensive set of neighborhood features related to cognitive health [4]. These neighborhood features could be classified into five categories: neighborhood deprivation, service facilities, food access, healthcare, and environmental hazards. A total of 21 individual measures were examined. All neighborhood measures were in the unit of counts per 1,000 population within a census tract and top-coded at the 99^th^ percentile to reduce the influence of high-leverage points. Detailed information of neighborhood variables is presented in Table 1.

**Table 1.** Neighborhood-Level Contextual Measures, NaNDA (2015) ^a^.

| Neighborhood Features | Indicators |
| --- | --- |
| Healthcare access | • Healthcare services including all ambulatory health care services, i.e., physicians and other health care providers.<br>• Nursing and residential care facilities<br>• Pharmacies and drug stores |
| Neighborhood service facilities | • Senior centers<br>• Arts sites and theaters<br>• Religious organizations<br>• Civic and social organizations<br>• Parks |
| Food stores/access | • Grocery stores<br>• Special food stores<br>• Fast food Restaurants<br>• Drinking places<br>• Coffee shops |
| Neighborhood deprivation indicators | • Proportion of population age 16+ unemployed<br>• Proportion of families with income below the federal poverty level<br>• Proportion of population with less than high school education<br>• Proportion of female headed families with children<br>• Proportion of black residents in the neighborhood<br>• Proportion of Hispanic residents in the neighborhood |
| Neighborhood environmental hazards | • Polluting sites within a census tract plus half-mile buffer<br>• Highways density: The density of highway stretches, derived through primary and secondary road segments |

<sup>a</sup> Neighborhood measures came from the National Neighborhood Data Archive (2015). All neighborhood measures were in the unit of counts per 1,000 population within a census tract.

#### Covariates

Demographic covariates included age (in years), gender (male = 0, female = 1), and race/ethnicity (non-Hispanic White = 1, non-Hispanic Black = 2, Hispanic = 3, or other = 4). Socioeconomic characteristics included educational attainment (less than high school = 1, high school or GED = 2, some college = 3, or college or above = 4), household income quartiles (lowest = 1, lower middle = 2, upper middle = 3, highest = 4), and working for payment (no = 0, yes = 1). We also included health-related covariates, including alcohol consumption (not drinking = 0 drinks/day, moderate drinking = men < 3 drinks/day or women < 2 drinks/day, excessive drinking = men ≥ 3 drinks/day or women ≥ 2 drinks/day). smoking status (never smoked =1, past smoker = 2, or current smoker = 3), and number of diagnosed chronic conditions (heart disease, lung disease, diabetes, cancer, arthritis, hypertension, and stroke). Disability status was assessed using the Activities of Daily Living (ADL) scale. The scale was constructed by summing binary indicators for difficulty in six basic activities: bathing, eating, dressing, walking across a room, getting in or out of bed, and using the toilet. We also included whether the participant was an apolipoprotein ε4 (APOE ε4) allele carrier. Geographic covariates were urbanicity (urban = 1, suburban = 2, or rural = 3) and census region (Northeast = 1, Midwest = 2, South = 3, or West = 4). Additional control variables, such as marital status, health insurance status, Medicare beneficiary status, and property ownership, were considered but omitted due to a lack of statistical significance.

### Statistical analyses

The analysis proceeded in two steps. First, we used Partitioning Around Medoids (PAM), an unsupervised machine-learning algorithm, to identify neighborhood typologies. PAM belongs to the family of partitioning clustering methods that group observations into a set of k clusters. PAM was chosen for its robustness to outliers, its ability to handle heterogeneous data types (both numerical and categorical data), and its capacity for irregular cluster geometries. This was crucial given the diversity and occasionally large values in our neighborhood measures. Euclidean distance was used as the dissimilarity metric for the clustering procedure. The optimal number of clusters was determined by combining the elbow method, visual inspection, and theoretical interpretability [34]. After obtaining the neighborhood cluster typologies, each cluster was subsequently assigned a descriptive label reflecting the distribution of the mean values of its constituent measures, aiming to best characterize its unique features. We then evaluated differences in sample characteristics across neighborhood typologies using the chi-square test or Kruskal-Wallis test, as appropriate.

Second, multilevel regression models were used to examine the relationship between neighborhood typologies and cognitive function. Given that individuals were nested within census tracts, we adopted a two-level model, with individuals as the first level and census tracts as the second level. We specified census tract-level random intercepts to account for the non-independence of observations within tracts. The covariance structure was set to identity to facilitate model estimation. These models were constructed using a stepwise covariate adjustment approach: Model 1 included neighborhood typologies and adjusted for demographic characteristics and geographic covariates, including age, gender, race, urbanity, and census region. Model 2 further adjusted for socioeconomic status, including educational attainment, household income, and working status. Model 3 additionally adjusted for chronic conditions, ADL, alcohol consumption, smoking status, and APOE ε4 allele carrier status. All models were weighted using HRS survey weights to account for complex study design and non-responses.

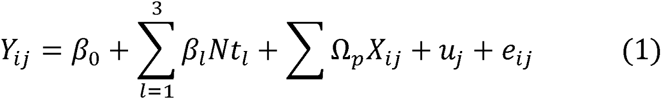

where *Y_ij_* represented the cognitive score for individual *i* nested within neighborhood *j*. The key explanatory variable *Nt_l_* was the identified neighborhood typologies. X*_ij_* represents covariates included in the model, such as demographic characteristics, geographic covariates, socioeconomic status, and health-related factors. β*_0_* was the intercept. The coefficients of interest are *β_l_*, for *l=1,…,3*, which quantified the relationship between living in different neighborhood typologies and cognitive outcomes, with the last neighborhood typology as the reference group. *Ω_p_* were the coefficients for covariates. u*_j_* represented the level-2 residual, which measured the difference between a specific neighborhood’s average cognitive score and the overall average cognitive score for all neighborhoods. e*_ij_* was the individual-level error term, which is assumed to have a bivariate normal distribution. Statistical tests were two-sided, with a significance level of *p < 0.05*. Data analyses were performed using RStudio version 4.0.2 [35] and STATA 17 [36].

Sensitivity analyses were conducted to evaluate the robustness of our findings. First, we included depression (assessed via the Center for Epidemiological Studies Depression [CES-D] scale) in the fully adjusted models to evaluate potential confounding due to its established association with cognitive function. Additionally, we further tested the robustness of our results by additionally adjusting for subjective perceptions of neighborhoods, specifically including perceived neighborhood disorder and neighborhood cohesion in the models.

## Results

The final analytic sample consisted of 6,480 community-dwelling older adults (Table 2). The mean age was 76 years (standard deviation [SD] = 7.5). 59.1% were female and nearly half (47.0%) had a college education. Over 72.4% of them were non-Hispanic White, 14.8% were non-Hispanic Black, 10.7% were Hispanic, and 2.2% were from other racial and ethnic groups. Eight percent of them reported being current smokers. Participants had an average of 2.4 chronic conditions (SD =1.3, range: 0-7). Approximately half of the participants lived in urban areas and 25.8% were APOE ε4 allele carriers. The mean cognitive score for the sample was 14.3 (SD=4.6, range: 0-27).

**Table 2.** Descriptive Statistics of the Study Sample (Health and Retirement, n = 6480) Variable.

| Variable | *n (%) / Mean (SD)<sup>a</sup>* |
| --- | --- |
| Cognitive function score (TICS), *mean (SD)* | 14.27 (4.60) |
| Female, *n (%)* | 3830 (59.10%) |
| Race, *n (%)* |  |
| Non-Hispanic White | 4688 (72.35%) |
| Non-Hispanic Black | 959 (14.80%) |
| Hispanic | 694 (10.71%) |
| Other | 139 (2.15%) |
| Age, *mean (SD)* | 75.96 (7.45) |
| Education, *n (%)* |  |
| Less than high school | 1149 (17.73%) |
| High school and GED | 2283 (35.23%) |
| Some college | 1536 (23.70%) |
| College and above | 1512 (23.33%) |
| Working for pay, *n (%)* |  |
| No | 5216 (80.49%) |
| Yes | 1264 (19.51%) |
| Household income quantiles, *n (%)* |  |
| Lowest | 1466 (22.62%) |
| Lower-middle | 2002 (30.90%) |
| Upper-middle | 1831 (28.26%) |
| Highest | 1181 (18.23%) |
| Alcohol consumptions, *n (%)* |  |
| Not drinking | 4215 (65.05%) |
| Moderate drinking | 1585 (24.46%) |
| Excessive drinking | 680 (10.49%) |
| Smoking, *n (%)* |  |
| Never smoked | 2953 (45.57%) |
| Past smokers | 3025 (46.68%) |
| Current smoker | 502 (7.75%) |
| Number of chronic conditions, *mean (SD)* | 2.43 (1.33) |
| ADL, *mean (SD)* | 0.41 (1.04) |
| Urbanity, *n (%)* |  |
| Urban | 3255 (50.23%) |
| Suburban | 1435 (22.15%) |
| Rural | 1790 (27.62%) |
| Census region, *n (%)* |  |
| Northeast | 993 (15.32%) |
| Mid-West | 1569 (24.21%) |
| South | 2705 (41.74%) |
| West | 1213 (18.72%) |
| APOE ε4 carrier, *n (%)* | 1671 (25.79%) |

Abbreviations: TICS = Telephone Interview for Cognitive Status; ADL = Activity of Daily Living;
APOE ε = ε apolipoprotein E.
<sup>a</sup> Frequency (n) with percentages (%) and mean with standard deviation (SD) are presented. These summary statistics are unweighted.

The Partitioning Around Medoids (PAM) was performed to identify distinct neighborhood typologies. The optimal number of clusters was primarily determined using the elbow method, which evaluates the reduction in the within-cluster sum of squares (WCSS) across candidate solutions. As shown in Appendix Figure S2, a four-cluster solution was selected at the point where the rate of WCSS decline slowed, indicating diminishing returns with additional clusters. Mean values of individual neighborhood measures for each cluster are presented in Appendix Figure S3 and Appendix Table S1. We identified four distinct clusters and labeled each cluster based on its corresponding characteristics. Cluster 1 was labelled as “**low deprivation, green neighborhood**” and was characterized by low neighborhood deprivation, sparse polluting sites, low highway density, very high park density, and a moderate density of neighborhood amenities, such as food stores, social organizations, and healthcare facilities (n = 1,957, 30.2%). Cluster 2 labeled “**Mid-SES, high hazard neighborhood,**” featured moderate socioeconomic deprivation, limited access to amenities and green space, and higher levels of environmental hazards (n = 2,451, 37.8%). Cluster 3 was designated the “**high-amenity neighborhood**” and characterized by moderate socioeconomic deprivation, but a high concentration of neighborhood amenities, services, and green space (n=1,068, 16.5%). Finally, Cluster 4 represented “**disadvantaged neighborhood,**” scoring highest on all deprivation indicators and exhibiting limited access to green space, essential services, and healthcare facilities, yet was marked by relatively low environmental hazards (n=1,004, 15.5%).

We present the descriptive statistics of sample characteristics across neighborhood typologies in Table 3. Significant differences were observed across 4 types of neighborhood clusters in sociodemographic characteristics, health behaviors, health status, and cognitive scores. Specifically, residents of the low deprivation, green neighborhoods (Cluster 1) were more likely to be older (M_age_ = 76.5, SD = 7.5), non-Hispanic White (87.9%), and college educated (56.9%). They also reported higher household income, with 24.7% belonging to the highest income quantile. This group was less likely to be current smokers (5.8%), had fewer chronic conditions (M=2.3, SD= 1.3) and were less likely to be APOE ε4 carriers (24.2%). The mean cognitive score for this group was 15.2 (SD=4.3). In contrast, individuals residing in Cluster 4 presented a profile characterized by multiple disadvantages. They were younger (M_age_ =74.6, SD=7.5) and had a lower household income, with only 7.1% in the highest income quantile. Only 28.5% had a college education or above. Racial and ethnic minority groups were overrepresented in this cluster (e.g., 42.1% Non-Hispanic Black; 35.9% Hispanic). Furthermore, they exhibited poorer health behaviors and status, including a higher prevalence of current smoking (11.3%), a higher average number of chronic conditions (M = 2.6, SD = 1.3), and a higher likelihood of being an APOE ε4 carrier (28.3%). They also reported a lower average cognitive score of 12.3 (SD = 4.7). Residents of the remaining two clusters exhibited comparable cognitive performance (M_cluster2_ = 14.4, SD = 4.6 vs. M_cluster3_ = 14.3, SD = 4.5). However, residents of the high-amenity neighborhoods (Cluster 3) demonstrated modestly higher socioeconomic status (e.g., higher education attainment and household income) yet poorer health behaviors (e.g., higher rates of smoking and excessive alcohol consumption), compared to residents of the Mid-SES, high hazard neighborhoods (Cluster 2).

**Table 3.** Sample Characteristics by Neighborhood Typologies.

|  | Neighborhood Typologies <sup>a, b</sup> |  |  |  | *p* <sup>c</sup> |
| --- | --- | --- | --- | --- | --- |
|  | Cluster 1 | Cluster 2 | Cluster 3 | Cluster 4 |  |
| n | 1957 (30.20%) | 2451 (37.82%) | 1068 (16.48%) | 1004 (15.49%) |  |
| Cognitive function score (TICS), *mean (SD)* | 15.15 (4.28) | 14.37 (4.60) | 14.28 (4.54) | 12.33 (4.67) | <0.001 |
| Female, *n (%)* | 1135 (58.00%) | 1415 (57.73%) | 649 (60.77%) | 631 (62.85%) | 0.02 |
| Race, *n (%)* |  |  |  |  | <0.001 |
| Non-Hispanic White | 1720 (87.89%) | 1994 (81.35%) | 779 (72.94%) | 195 (19.42%) |  |
| Non-Hispanic Black | 100 (5.11%) | 269 (10.98%) | 167 (15.64%) | 423 (42.13%) |  |
| Hispanic | 96 (4.91%) | 132 (5.39%) | 106 (9.93%) | 360 (35.86%) |  |
| Other | 41 (2.10%) | 56 (2.28%) | 16 (1.50%) | 26 (2.59%) |  |
| Age, *mean (SD)* | 76.45 (7.48) | 75.96 (7.32) | 76.38 (7.51) | 74.57 (7.49) | <0.001 |
| Education, *n (%)* |  |  |  |  | <0.001 |
| Less than high school | 183 (9.35%) | 374 (15.26%) | 176 (16.48%) | 416 (41.43%) |  |
| High school and GED | 660 (33.73%) | 980 (39.98%) | 341 (31.93%) | 302 (30.08%) |  |
| Some college | 497 (25.40%) | 570 (23.26%) | 274 (25.66%) | 195 (19.42%) |  |
| College and above | 617 (31.53%) | 527 (21.50%) | 277 (25.94%) | 91 (9.06%) |  |
| Working for pay, *n (%)* |  |  |  |  | 0.63 |
| No | 1557 (79.56%) | 1983 (80.91%) | 860 (80.52%) | 816 (81.27%) |  |
| Yes | 400 (20.44%) | 468 (19.09%) | 208 (19.48%) | 188 (18.73%) |  |
| Household income quantiles, *n (%)* |  |  |  |  | <0.001 |
| Lowest | 275 (14.05%) | 513 (20.93%) | 251 (23.50%) | 427 (42.53%) |  |
| Lower-middle | 532 (27.18%) | 831 (33.90%) | 320 (29.96%) | 319 (31.77%) |  |
| Upper-middle | 667 (34.08%) | 704 (28.72%) | 273 (25.56%) | 187 (18.63%) |  |
| Highest | 483 (24.68%) | 403 (16.44%) | 224 (20.97%) | 71 (7.07%) |  |
| Alcohol consumptions, *n (%)* |  |  |  |  | <0.001 |
| Not drinking | 1091 (55.75%) | 1663 (67.85%) | 688 (64.42%) | 773 (76.99%) |  |
| Moderate drinking | 638 (32.60%) | 550 (22.44%) | 268 (25.09%) | 129 (12.85%) |  |
| Excessive drinking | 228 (11.65%) | 238 (9.71%) | 112 (10.49%) | 102 (10.16%) |  |
| Smoking, *n (%)* |  |  |  |  | <0.001 |
| Never smoked | 901 (46.04%) | 1100 (44.88%) | 490 (45.88%) | 462 (46.02%) |  |
| Past smokers | 943 (48.19%) | 1163 (47.45%) | 490 (45.88%) | 429 (42.73%) |  |
| Current smoker | 113 (5.77%) | 188 (7.67%) | 88 (8.24%) | 113 (11.25%) |  |
| Number of chronic conditions, *mean (SD)* | 2.31 (1.31) | 2.49 (1.33) | 2.38 (1.31) | 2.58 (1.32) | <0.001 |
| ADL, *mean (SD)* | 0.29 (0.85) | 0.39 (1.00) | 0.42 (1.05) | 0.66 (1.34) | <0.001 |
| Urbanity, *n (%)* |  |  |  |  | <0.001 |
| Urban | 1206 (61.62%) | 893 (36.43%) | 560 (52.43%) | 596 (59.36%) |  |
| Suburban | 481 (24.58%) | 510 (20.81%) | 169 (15.82%) | 275 (27.39%) |  |
| Rural | 270 (13.80%) | 1048 (42.76%) | 339 (31.74%) | 133 (13.25%) |  |
| Census region, *n (%)* |  |  |  |  | <0.001 |
| Northeast | 390 (19.93%) | 299 (12.20%) | 146 (13.67%) | 158 (15.74%) |  |
| Mid-West | 519 (26.52%) | 657 (26.81%) | 270 (25.28%) | 123 (12.25%) |  |
| South | 585 (29.89%) | 1149 (46.88%) | 457 (42.79%) | 514 (51.20%) |  |
| West | 463 (23.66%) | 346 (14.12%) | 195 (18.26%) | 209 (20.82%) |  |
| APOE ε4 carrier, *n (%)* | 473 (24.17%) | 622 (25.38%) | 292 (27.34%) | 284 (28.29%) | <0.001 |

Abbreviations: TICS = Telephone Interview for Cognitive Status; ADL = Activity of Daily Living;
APOE ε = ε apolipoprotein E.
<sup>a</sup> Frequencies with percentages (%) for categorical variables and means with standard deviations (*SD*) for continuous variables are presented.
<sup>b</sup> Neighborhood Typologies: Cluster 1: Low deprivation, green neighborhood; Cluster 2: Mid-SES, high hazard neighborhood; Cluster 3: High-amenity neighborhood; Cluster 4: Disadvantaged neighborhood.
<sup>c</sup> $\chi^2$ test or Kruskal Wallis test were performed to compare statistical differences.

Results from multilevel models are presented in Table 4 (full models are available in Appendix Table S2). Compared to those living in disadvantaged neighborhoods (Cluster 4), residents of low deprivation, green neighborhoods (Cluster 1: *β* = 5.2, *95% CI*: 3.8 – 6.7), mid-SES, high-hazard neighborhoods (Cluster 2: *β* = 5.2, *95% CI*: 1.1 – 9.4), and high-amenity neighborhoods (Cluster 3: *β* = 5.5, *95% CI*: 2.3 – 8.7) demonstrated significantly higher cognitive function scores, adjusting for demographic characteristics and area of residence (Model 1). Residents of high-amenity neighborhoods (Cluster 3) continued to show significantly higher cognitive function scores (*β* = 4.7, *95% CI*: 1.9 – 7.4) when socioeconomic characteristics were included (Model 2). The associations were attenuated and became nonsignificant for those in low deprivation, green neighborhoods (Cluster 1: *β* = 1.5, *95% CI*: -0.7 – 3.8) and those in mid-SES, high-hazard neighborhoods (Cluster 2: *β* = 3.0, *95% CI*: -0.5 – 6.4) (Model 2). In the fully adjusted model (Model 3), residence in high-amenity neighborhoods (Cluster 3) remained significantly associated with higher cognitive function scores, though with some attenuation of the estimate (*β* = 3.9, *95% CI*: 1.2 – 6.5). Associations for the other neighborhood clusters remained statistically nonsignificant in the fully adjusted model.

**Table 4.** Multilevel Regression Estimating the Association between Neighborhood Typologies and Cognitive Function.

|  | Model 1<sup>a</sup> | Model 2<sup>b</sup> | Model 3<sup>c</sup> |
| --- | --- | --- | --- |
| | $\beta$ [95% *CI*]<sup>d</sup> | $\beta$ [95% *CI*]<sup>d</sup> | $\beta$ [95% *CI*]<sup>d</sup> |
| Neighborhood (ref. Cluster 4:<br>Disadvantaged neighborhood) |  |  |  |
| Cluster 1: Low deprivation, green<br>neighborhood | 5.24***<br>[3.75, 6.73] | 1.53<br>[-0.73, 3.78] | 1.10<br>[-1.03, 3.24] |
| Cluster 2: Mid-SES, high-hazard<br>neighborhood | 5.23*<br>[1.06, 9.40] | 2.96<br>[-0.51, 6.42] | 2.53<br>[-0.71, 5.78] |
| Cluster 3: High-amenity neighborhood | 5.46**<br>[2.25, 8.67] | 4.69**<br>[1.94, 7.44] | 3.85**<br>[1.23, 6.47] |
| Intercept | 9.84**<br>[7.70, 11.99] | 22.03**<br>[18.65, 25.41] | 23.14**<br>[19.85, 26.43] |
| Variance (intercept) | 17.66<br>[16.76, 18.62] | 16.05<br>[15.21, 16.92] | 14.83<br>[14.03, 15.67] |
| Variance (residual) | 7.61<br>[7.11, 8.16] | 6.03<br>[5.64, 6.45] | 5.93<br>[5.55, 6.34] |

Abbreviations: CI = confidence interval.
<sup>a</sup> Model 1 adjusted for age, gender, race, urbanity, and census region.
<sup>b</sup> Model 2 adjusted for education, income, working status and model 1 covariates.
<sup>c</sup> Model 3 adjusted for comorbidity, Activity of Daily Living (ADL), alcohol consumption, smoking, APOE ε4 carrier, and model 2 covariates.
<sup>d</sup> Regression coefficients and 95% confidence intervals were reported.
\*\* $p < 0.01$ , \* $p < 0.05$ .

We conducted sensitivity analyses to assess the robustness of our findings. Additionally accounting for depression, perceived neighborhood disorder, or perceived neighborhood cohesion resulted in similar findings to the regression estimates presented in the main analysis (Appendix Table S3).

## Discussion

This study extended the scholarship of the neighborhood exposome by moving beyond individual contextual risk factors to consider the “totality of neighborhood environments.” While some studies have employed composite measures of neighborhood socioeconomic disadvantage or deprivation indices, these indices fail to capture the multifaceted combination of amenities, services, and physical environments that may collectively shape cognitive health in later life. By applying a clustering approach to a large, nationally representative sample of older adults, our analysis yielded two major findings. First, we identified four neighborhood typologies, capturing a spectrum of neighborhoods differentiated by their composite profiles of protective and risk factors. Second, we found that older adults residing in resourceful neighborhoods characterized by extensive facilities and cognitively stimulating services demonstrated better cognitive function compared to those in disadvantaged, resource-scarce neighborhoods.

The identified typologies illustrated the complexity of neighborhood contexts and the way socioeconomic, structural, and environmental factors intersect at the neighborhood level. For instance, while the low-deprivation, green neighborhood cluster (Cluster 1) aligns with established findings that affluent neighborhoods tend to have fewer hazards and greater access to resources [37], we also identified a cluster characterized by moderate socioeconomic deprivation with a high density of amenities and green space (Cluster 3). Social services and facilities are likely to concentrate in areas with a high need for social support among older adults. A recent study found that the density of social care organizations is higher in less affluent neighborhoods with a greater number of older adults [38], which corresponds with the characteristics of Cluster 3 found in our analysis. At the same time, a neighborhood cluster marked by moderate deprivation and elevated environmental hazards also emerged (Cluster 2), suggesting that residents in these areas may experience overlapping social and environmental risks. Furthermore, Cluster 4 was characterized by pronounced socioeconomic disadvantage coupled with low hazards. Even with fewer toxic exposures, the absence of protective resources, such as healthcare and green space, may still pose long-term health influences on older residents. While research on neighborhood typologies remains scarce and largely exploratory, these patterns resonate with emerging evidence. For example, Humphrey et al. [39] similarly identified four neighborhood clusters within a community-based urban sample, noting that moderate-poverty neighborhoods were characterized by poor or moderate physical environments, whereas wealthy neighborhoods benefited from better physical environments and fewer hazards. Taken together, our identified neighborhood typologies reveal that neighborhoods are complex systems where structural disadvantages and protective resources may coexist in varied combinations, highlighting the importance of a multidimensional framework to capture the health implications of neighborhood environments.

Our findings revealed that older adults residing in high-amenity neighborhoods (Cluster 3) exhibited significantly better cognitive function than those living in neighborhoods characterized by socioeconomic disadvantage and limited access to amenities and resources (Cluster 4). This pattern is consistent with prior cross-national research demonstrating the influence of multidimensional neighborhood characteristics on healthy aging. For example, Wong et al. [40], drawing on data from 33 European countries, found that access to healthcare services, amenities, and green spaces were important predictors of optimal aging outcomes. Echoing the ecological framework of place [24,41], our findings also highlighted that neighborhoods with greater access to a constellation of amenities such as healthcare, social services, and civic organizations were linked to better cognitive function for the residents. The cognitive advantages observed in these high-amenity communities may stem from several interconnected mechanisms. Neighborhoods rich in civic and social organizations provide abundant opportunities for cognitively stimulating activities. Access to arts venues, museums, and recreation centers can encourage participation in complex, challenging, and interactive activities that activate multiple cognitive domains and stimulate neuroplasticity [4]. Meanwhile, these settings often encourage physical activity (e.g., group exercises or community volunteering), social interactions, and network building, which in turn support physical health (e.g., cardiovascular health) and reduce stress and inflammation [19,42–44]. The routine engagement in this blend of stimulating experiences is fundamental to enhancing neural efficiency and building a cognitive reserve that buffers against age-related cognitive decline.

Notably, we did not find significant differences in cognitive function between older adults residing in moderate-SES, high hazard neighborhoods (Cluster 2) and those in disadvantaged, resource-scarce neighborhoods (Cluster 4). The nonsignificant finding may reflect the complex interplay of protective and risk factors that operate simultaneously in shaping cognitive health. While a moderate socioeconomic position may be a protective factor, Cluster 2 was characterized by a scarcity of cognitively stimulating resources, such as art organizations, community centers, and parks, which are established resources supporting healthy aging and cognitive function [10,40]. Importantly, Cluster 2 also exhibited the highest levels of environmental hazards, including dense highways and many polluting sites. These risk factors have been linked to an increased risk of neuroinflammation, accelerated cognitive decline, and the accumulation of Alzheimer’s-related pathologies among older adults [8,17]. Consistent with a prior national cohort study, Finlay (2022) found that highway exposure was among the strongest predictors of geographic disparities in cognitive function. Proximity to highways may reduce walkability, heighten safety concerns from fast-moving traffic, increase exposure to air pollution, and limit access to everyday destinations. These factors may further restrict opportunities for physical activity and social engagement. Collectively, these findings align with a growing body of literature suggesting that environmental toxins and hazards may constitute major independent risk factors for late-life cognitive health.

It is noteworthy that cognitive function among older adults in low deprivation, green neighborhoods (Cluster 1) was comparable to those in disadvantaged neighborhoods (Cluster 4), despite prior evidence supporting a cognitive benefit from living in socioeconomically advantageous neighborhoods [10,11]. A possible explanation involves residential selection [45]. Our regression analysis showed that residing in low-deprivation, green neighborhoods was significantly associated with better cognitive function in unadjusted models. This association was attenuated to non-significance after adjusting for individual-level socioeconomic characteristics and health-related factors. This finding may suggest that the cognitive advantages observed among residents of Cluster 1 might largely be explained by compositional effects. That is, individuals with greater socioeconomic resources and better health profiles may be more likely to reside in affluent communities, rather than by a direct contextual effect of the neighborhood environment itself. Since individuals do not randomly occupy neighborhoods but instead choose or are constrained into residences based on socioeconomic, cultural, and health-related factors, the observed neighborhood effect in the initial model might indicate a systematic sorting process [19,46]. Nonetheless, this finding does not rule out meaningful contextual contributions, as we identified significant associations in the high-amenity neighborhood (Cluster 3) even after accounting for individual-level factors. Therefore, future studies could employ designs capable of addressing selection processes to better capture causal pathways through which neighborhood contextual features shape cognitive health.

Several limitations should be noted. First, while machine learning clustering methods offer a powerful tool for identifying complex patterns, the approach can introduce uncertainty given the “black box” nature of the algorithm. We chose a four-cluster solution for this analysis based on the elbow method and the distribution of observations across clusters. We also considered a five-cluster solution, but it included a cluster with a small number of observations, which would have resulted in cells with insufficient cases for subsequent analyses. Moreover, because this study relied on a sample from the Health and Retirement Study, replication with a wider range of community and clinical samples is needed to test the external validity and generalizability of the identified neighborhood typologies. Second, while our typologies incorporated key dimensions including socioeconomic status, environmental hazards, and access to amenities and services, other important neighborhood features, such as crime, safety, or transportation infrastructure, were not captured in this analysis. Future research should consider a broader array of neighborhood features to provide a more comprehensive understanding of environmental influences on cognitive aging. Third, we were not able to assess changes in neighborhood context and cognition over time due to the cross-sectional nature of this study. Future research employing longitudinal data is crucial to unravel the causal relationship between neighborhood typologies and cognitive health.

## Conclusions

Our study advances understanding of multidimensional neighborhood features in relation to cognitive function among community-dwelling older Americans. We identified four distinct neighborhood typologies. Our findings also underscore significant disparities in cognitive outcomes among older residents across neighborhood contexts, with resource-rich neighborhoods characterized by abundant amenities and cognitively stimulating facilities demonstrating better cognitive function. The implications of our findings lie in the importance of addressing modifiable neighborhood factors, particularly those related to amenity availability and environmental hazards. Tailored community-level interventions could be pivotal in promoting cognitive health among older adults within diverse neighborhood contexts.

## Data Availability

Data for this research can be accessed via HRS (https://hrs.isr.umich.edu/data-products) and NaNDA(https://nanda.isr.umich.edu/data/) websites.

## Supplemental Materials

**Appendix Figure S1.**
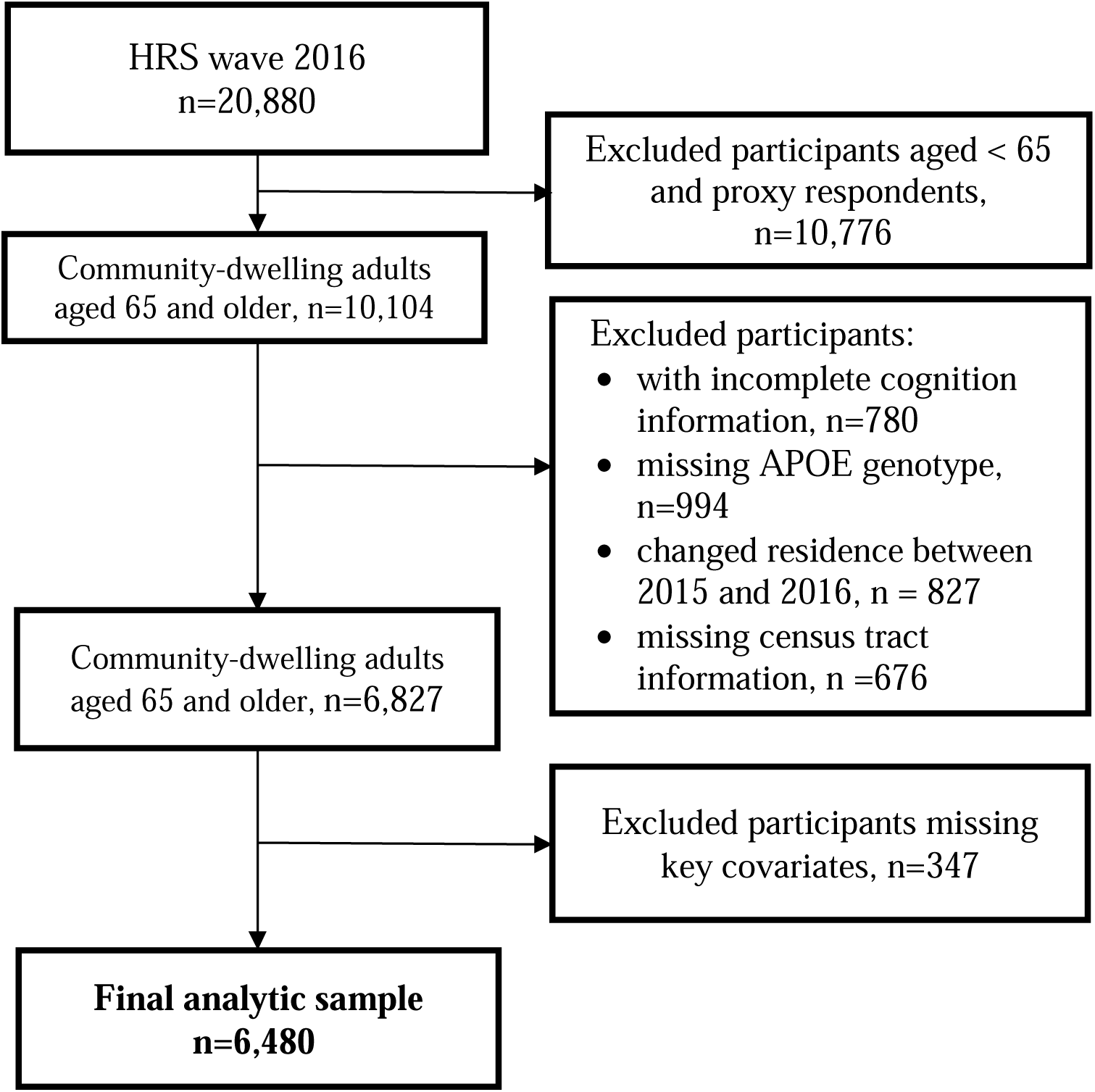
Sample selection flow chart

**Appendix Figure S2.**
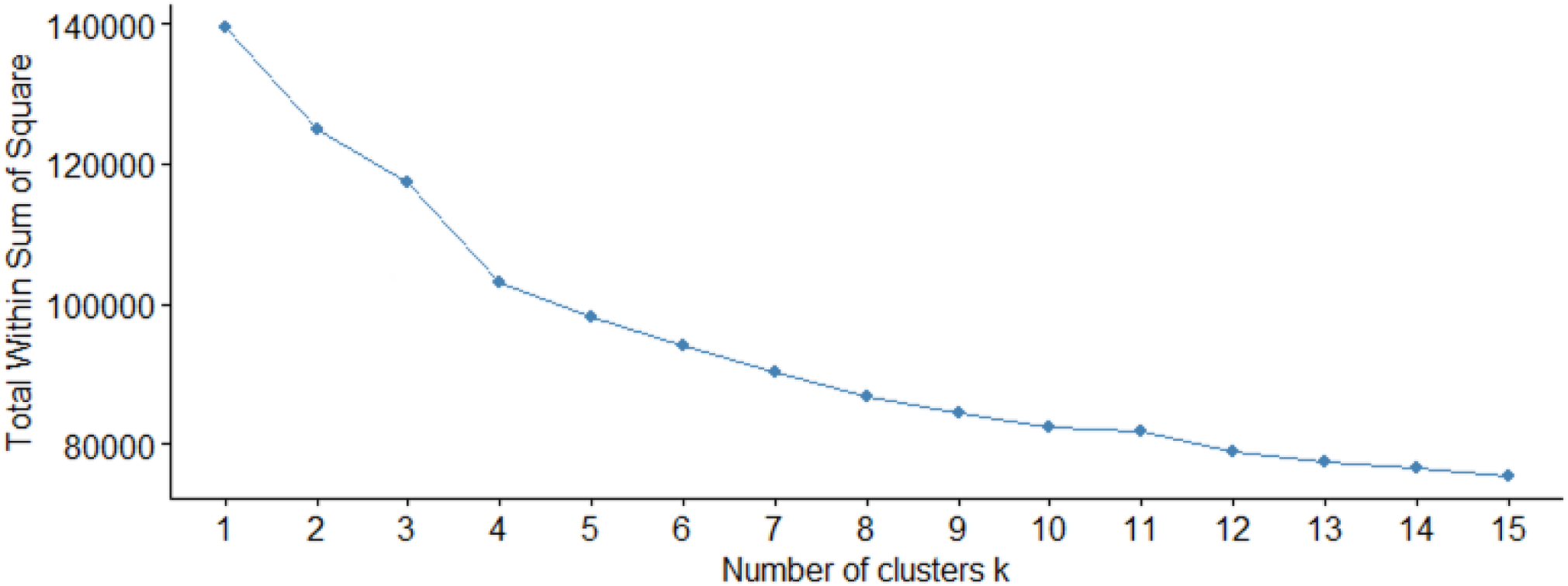
Total within sum of square plot on number of clusters. The graph shows the within-cluster-sum-of-square (WCSS) values on the y-axis corresponding to the different values of clusters K (on the x-axis). The optimal K value is the point at which the graph forms an elbow or where the rate of WCSS decline slowed, indicating diminishing returns with additional clusters.

**Appendix Figure S3.**
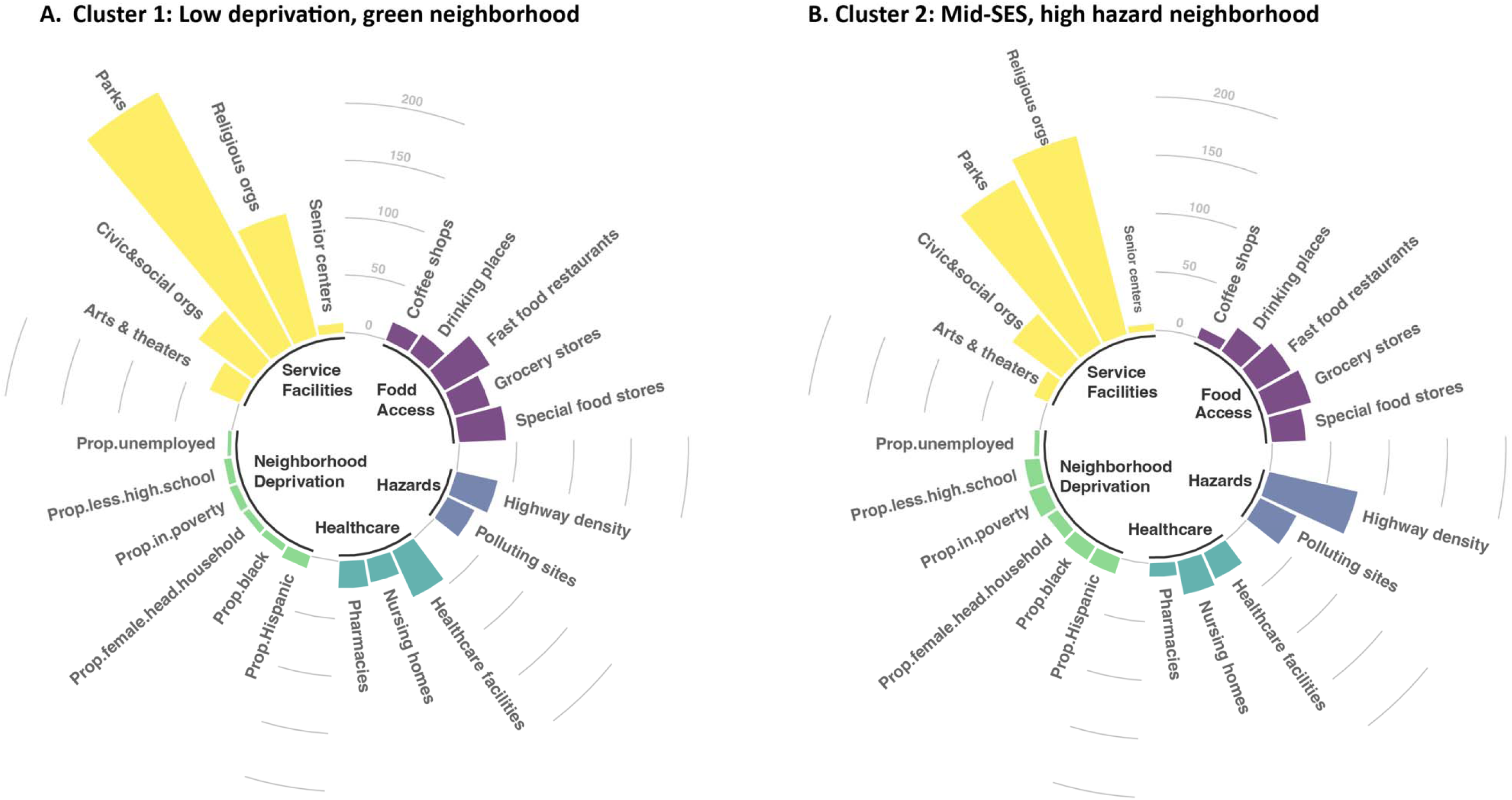
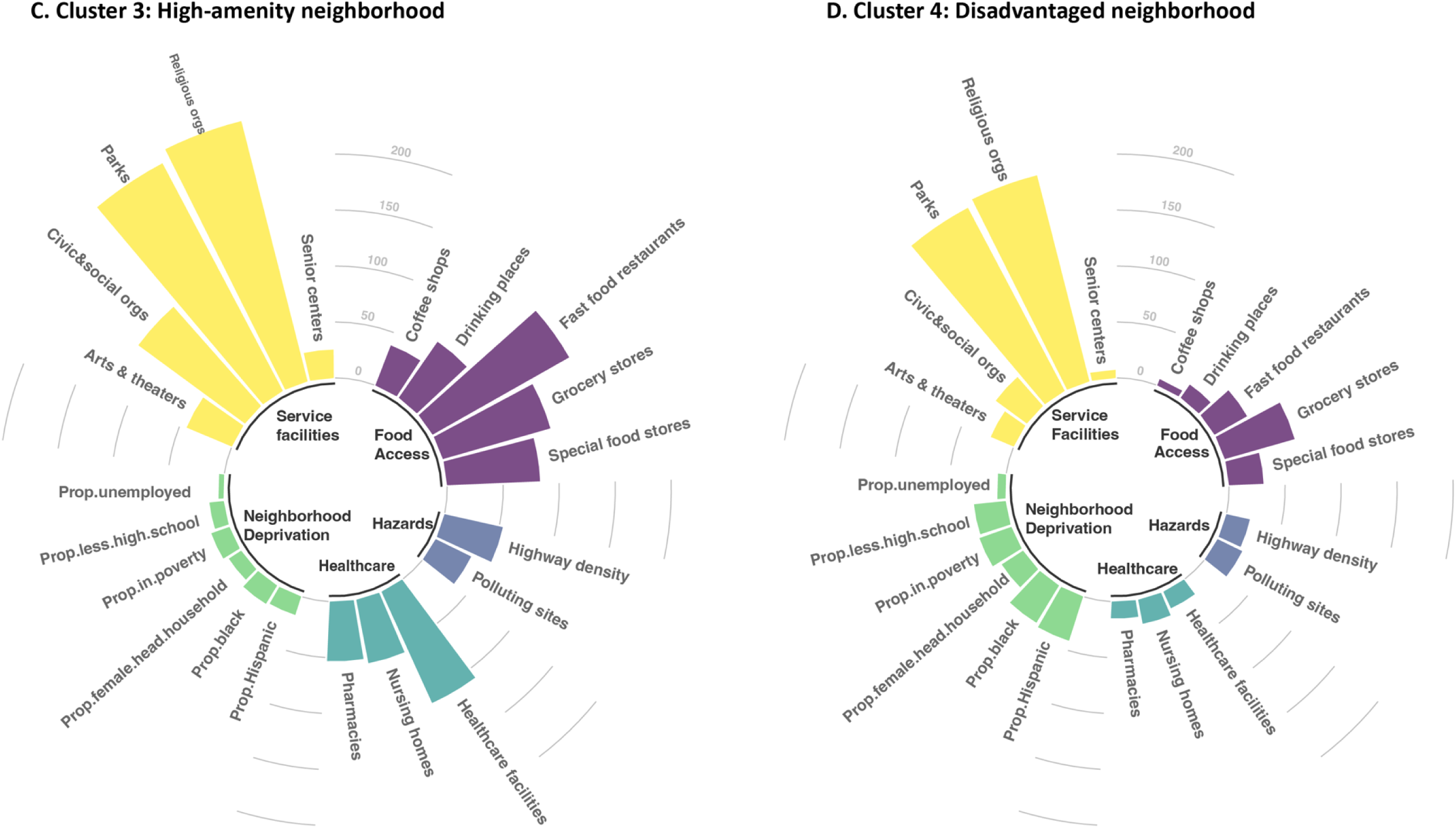
The distribution of neighborhood-level measures for neighborhood typologies Note: Mean values of individual neighborhood measures for each cluster are presented. All neighborhood measures were in the unit of counts per 1,000 population within a census tract.

**Appendix Table S1.**
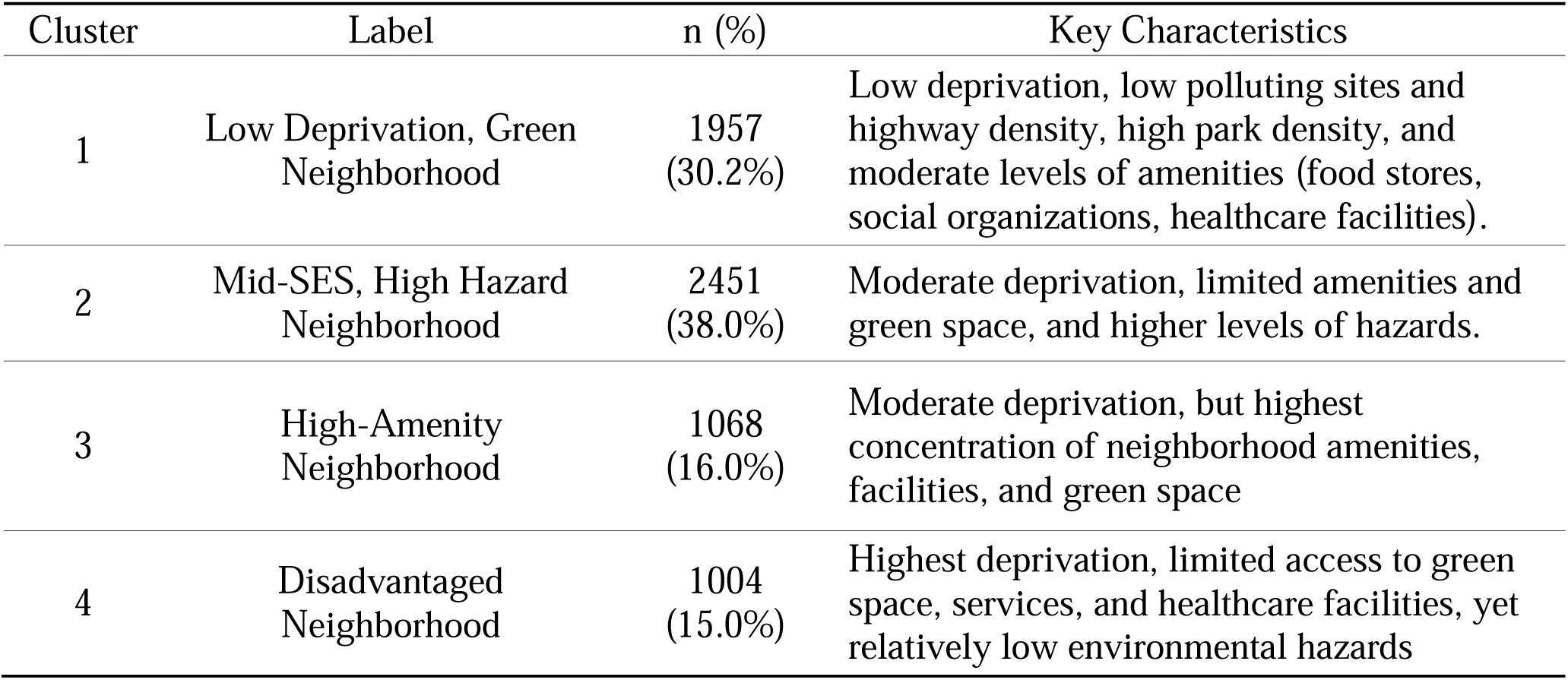
Summary of identified neighborhood typologies.

**Appendix Table S2.**
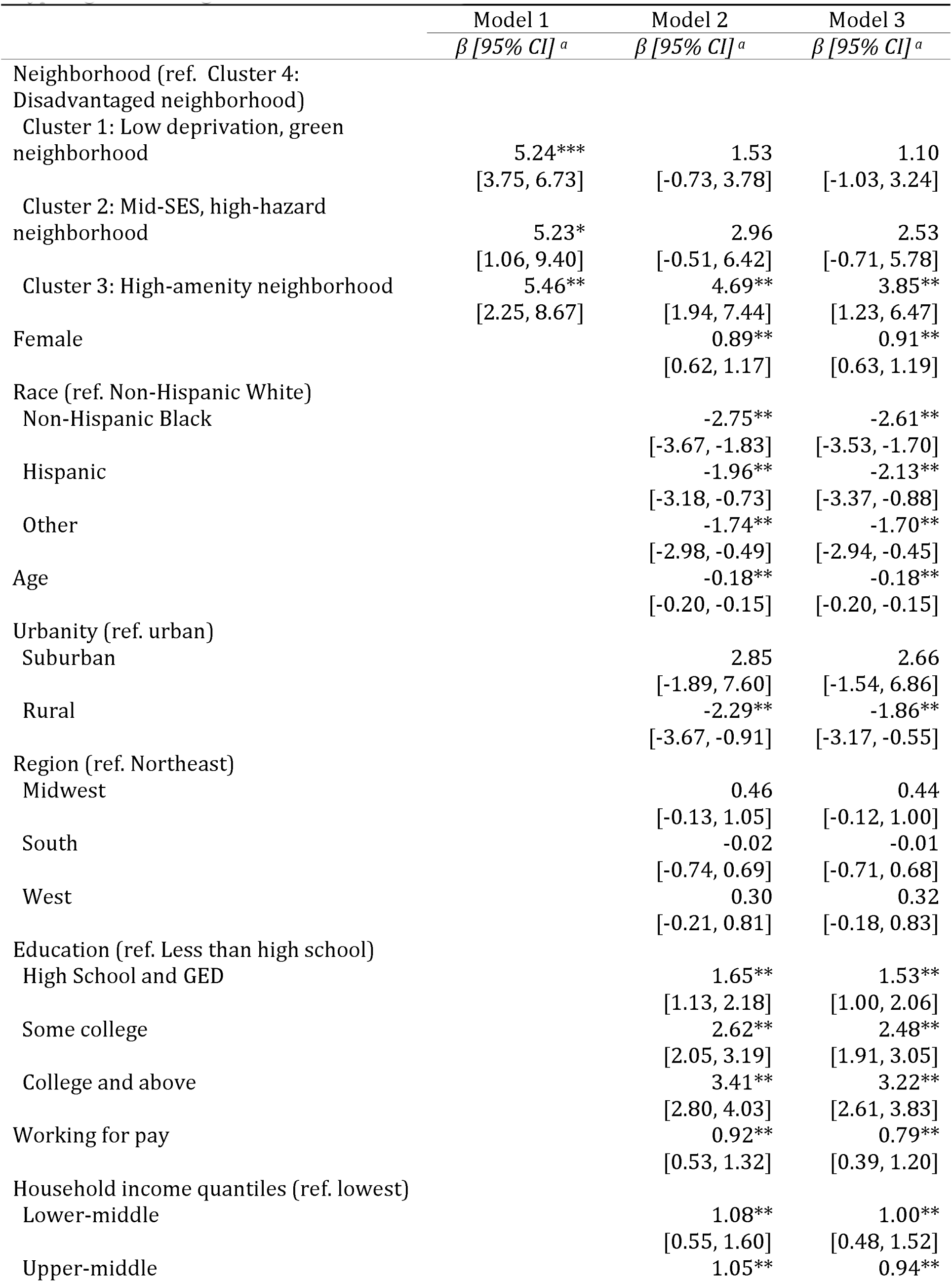
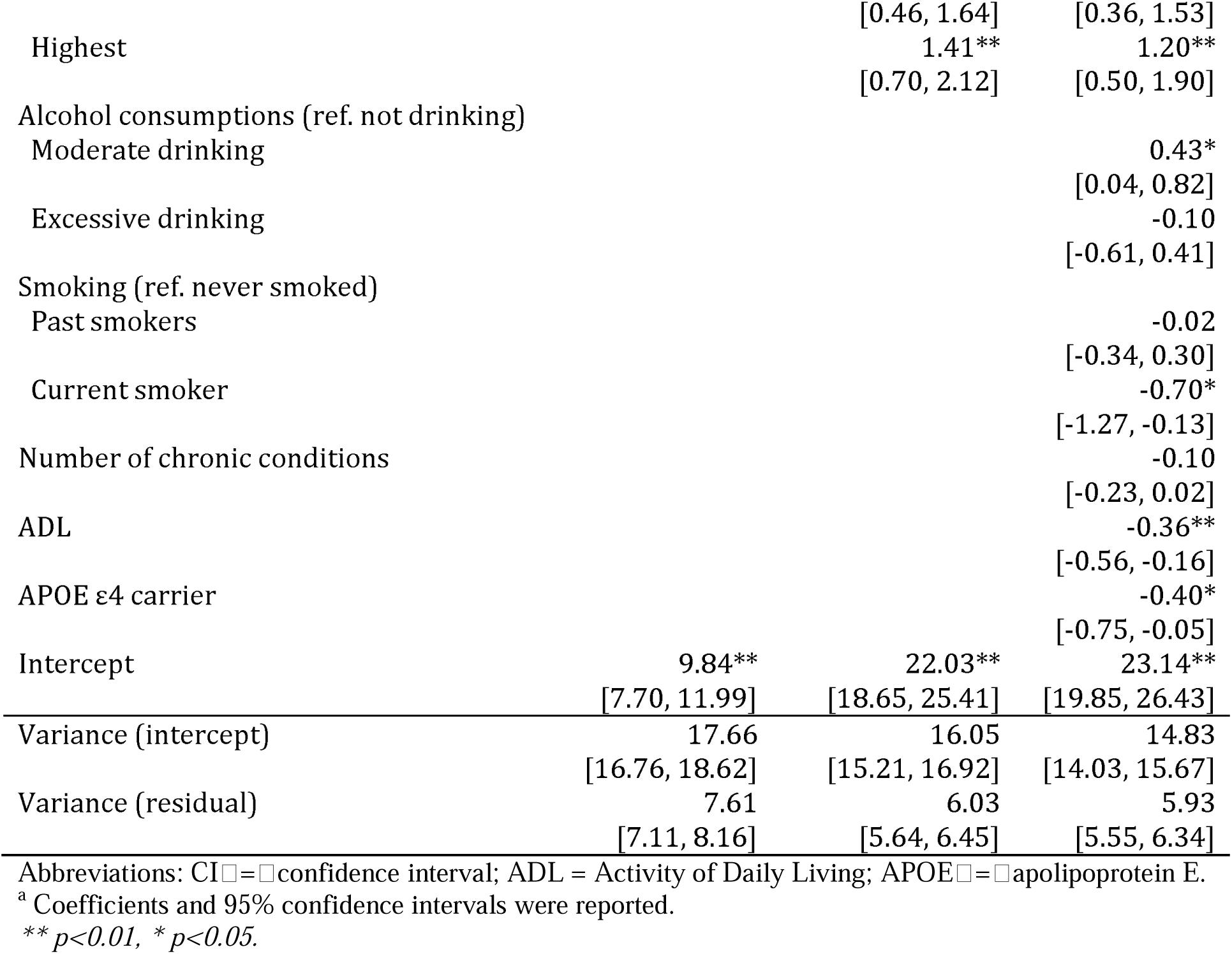
Multilevel Regression Estimating the Association between Neighborhood Typologies and Cognitive Function.

**Appendix Table S3.** Multilevel Regression Estimating Associations between Neighborhood Typologies and Cognitive Function Adjusting for Additional Covariates.

|  | Adjusted estimate (95% CI) <sup>a</sup> |  |  |
| --- | --- | --- | --- |
|  | Adjusting for<br>covariates and<br>depression | Adjusting for<br>covariates and<br>neighborhood<br>disorder | Adjusting for<br>covariates<br>and<br>neighborhood<br>cohesion |
| Neighborhood (ref. Cluster 4:<br>Disadvantaged neighborhood) |  |  |  |
| Cluster 1: Low deprivation, green<br>neighborhood | 1.12<br>[-1.03, 3.27] | 1.27<br>[-1.74, 4.28] | 1.78<br>[-1.24, 4.80] |
| Cluster 2: Mid-SES, high-hazard<br>neighborhood | 2.54<br>[-0.79, 5.87] | 2.77<br>[-0.85, 6.40] | 3.24<br>[-0.51, 6.99] |
| Cluster 3: High-amenity<br>neighborhood | 3.77**<br>[1.16, 6.38] | 4.88**<br>[1.30, 8.46] | 5.21**<br>[1.62, 8.82] |

Abbreviations: CI = confidence interval
<sup>a</sup> All models adjusted for age, gender, race, urbanity, census region, education, income, working status, comorbidity, Activity of Daily Living (ADL), alcohol consumption, smoking, APOE ε4 carrier.
\*\* $p < 0.01$ , \* $p < 0.05$ .

## References

1. Manly JJ, Jones RN, Langa KM, Ryan LH, Levine DA, McCammon R, et al. Estimating the Prevalence of Dementia and Mild Cognitive Impairment in the US: The 2016 Health and Retirement Study Harmonized Cognitive Assessment Protocol Project. JAMA Neurol. 2022;79: 1242–1249. doi:10.1001/jamaneurol.2022.3543

2. Yu J, Cudjoe TKM, Mathis WS, Chen X. Uncovering the Biological Toll of Neighborhood Physical Disorder: Links to Metabolic and Inflammatory Biomarkers in Older Adults. medRxiv; 2024. p. 2024.12.23.24319571. doi:10.1101/2024.12.23.24319571

3. Besser L, McDonald NC, Song Y, Kukull WA, Rodriguez DA. Neighborhood Environment and Cognition in Older Adults: A Systematic Review. American Journal of Preventive Medicine. 2017;53: 241–251. doi:10.1016/j.amepre.2017.02.013

4. Finlay J, Esposito M, Langa KM, Judd S, Clarke P. Cognability: An Ecological Theory of neighborhoods and cognitive aging. Social Science & Medicine. 2022;309: 115220. doi:10.1016/j.socscimed.2022.115220

5. Lee H, Waite LJ. Cognition in Context: The Role of Objective and Subjective Measures of Neighborhood and Household in Cognitive Functioning in Later Life. The Gerontologist. 2018;58: 159–169. doi:10.1093/geront/gnx050

6. Yang T-C, Kim S, Choi SE, Halloway S, Mitchell UA, Shaw BA. Neighborhood Features and Cognitive Function: Moderating Roles of Individual Socioeconomic Status. American Journal of Preventive Medicine. 2024;66: 454–462. doi:10.1016/j.amepre.2023.10.012

7. Godina SL, Rosso AL, Hirsch JA, Besser LM, Lovasi GS, Donovan GH, et al. Neighborhood greenspace and cognition: The cardiovascular health study. Health & Place. 2023;79: 102960. doi:10.1016/j.healthplace.2022.102960

8. Kim B, Samuel LJ, Thorpe Jr RJ, Crews DC, Szanton SL. Food Insecurity and Cognitive Trajectories in Community-Dwelling Medicare Beneficiaries 65 Years and Older. JAMA Network Open. 2023;6: e234674–e234674. doi:10.1001/jamanetworkopen.2023.4674

9. Rosso AL, Harding AB, Clarke P, Studenski SA, Rosano C. Associations of Neighborhood Walkability and Walking Behaviors by Cognitive Trajectory in Older Adults. Gerontologist. 2021;61: 1053–1061. doi:10.1093/geront/gnab005

10. Clarke P, Weuve J, Barnes L, Evans DA, Mendes de Leon CF. Cognitive decline and the neighborhood environment. Annals of Epidemiology. 2015;25: 849–854. doi:10.1016/j.annepidem.2015.07.001

11. Dintica CS, Bahorik A, Xia F, Kind A, Yaffe K. Dementia Risk and Disadvantaged Neighborhoods. JAMA Neurology. 2023;80: 903–909. doi:10.1001/jamaneurol.2023.2120

12. Wong R, Wang Y. Role of Neighborhood Physical Disorder and Social Cohesion on Racial and Ethnic Disparities in Dementia Risk. J Aging Health. 2022;34: 1178–1187. doi:10.1177/08982643221101352

13. Rosso AL, Grubesic TH, Auchincloss AH, Tabb LP, Michael YL. Neighborhood Amenities and Mobility in Older Adults. American Journal of Epidemiology. 2013;178: 761–769. doi:10.1093/aje/kwt032

14. Levasseur M, Gauvin L, Richard L, Kestens Y, Daniel M, Payette H. Associations Between Perceived Proximity to Neighborhood Resources, Disability, and Social Participation Among Community-Dwelling Older Adults: Results From the VoisiNuAge Study. Archives of Physical Medicine and Rehabilitation. 2011;92: 1979–1986. 10.1016/j.apmr.2011.06.035

15. Bourassa KJ, Memel M, Woolverton C, Sbarra DA. Social participation predicts cognitive functioning in aging adults over time: comparisons with physical health, depression, and physical activity. Aging & Mental Health. 2017;21: 133–146. doi:10.1080/13607863.2015.1081152

16. Besser L, Galvin JE, Rodriguez D, Seeman T, Kukull W, Rapp SR, et al. Associations between neighborhood built environment and cognition vary by apolipoprotein E genotype: Multi-Ethnic Study of Atherosclerosis. Health & Place. 2019;60: 102188. doi:10.1016/j.healthplace.2019.102188

17. Ailshire J, Karraker A, Clarke P. Neighborhood social stressors, fine particulate matter air pollution, and cognitive function among older U.S. adults. Social Science & Medicine. 2017;172: 56–63. 10.1016/j.socscimed.2016.11.019

18. Gessa GD, Bloomberg M, So R, Scholes S, Byrne T, Lee J, et al. Cognitive Performance and Long-term Exposure to Outdoor Air Pollution: Findings From the Harmonized Cognitive Assessment Protocol Substudy of the English Longitudinal Study of Ageing (ELSA-HCAP). The Journals of Gerontology: Series A. 2025;80: glaf060. doi:10.1093/gerona/glaf060

19. Glymour MM, Mujahid M, Wu Q, White K, Tchetgen Tchetgen EJ. Neighborhood Disadvantage and Self-Assessed Health, Disability, and Depressive Symptoms: Longitudinal Results From the Health and Retirement Study. Annals of Epidemiology. 2010;20: 856–861. 10.1016/j.annepidem.2010.08.003

20. Padeiro M, de São José J, Amado C, Sousa L, Roma Oliveira C, Esteves A, et al. Neighborhood Attributes and Well-Being Among Older Adults in Urban Areas: A Mixed-Methods Systematic Review. Research on Aging. 2021;44: 351–368. doi:10.1177/0164027521999980

21. Rosso AL, Flatt JD, Carlson MC, Lovasi GS, Rosano C, Brown AF, et al. Neighborhood Socioeconomic Status and Cognitive Function in Late Life. American Journal of Epidemiology. 2016;183: 1088–1097. doi:10.1093/aje/kwv337

22. Mejía ST, Ryan LH, Gonzalez R, Smith J. Successful Aging as the Intersection of Individual Resources, Age, Environment, and Experiences of Well-being in Daily Activities. The Journals of Gerontology: Series B. 2017;72: 279–289. doi:10.1093/geronb/gbw148

23. Flores M, Ruiz JM, Butler EA, Sbarra DA. Hispanic Ethnic Density May Be Protective for Older Black/African American and Non-Hispanic White Populations for Some Health Conditions: An Exploration of Support and Neighborhood Mechanisms. Annals of Behavioral Medicine. 2022;56: 21–34. doi:10.1093/abm/kaab014

24. Moore KD. An Ecological Framework of Place: Situating Environmental Gerontology within a Life Course Perspective. The International Journal of Aging and Human Development. 2014;79: 183–209. doi:10.2190/AG.79.3.a

25. Hartig T, Mitchell R, de Vries S, Frumkin H. Nature and Health. Annual Review of Public Health. 2014;35: 207–228. doi:10.1146/annurev-publhealth-032013-182443

26. Lawton MP, Nahemow L. Ecology and the aging process. The psychology of adult development and aging. Washington, DC, US: American Psychological Association; 1973. pp. 619–674. doi:10.1037/10044-020

27. Migeot J, Pina-Escudero SD, Hernandez H, Gonzalez-Gomez R, Legaz A, Fittipaldi S, et al. Social exposome and brain health outcomes of dementia across Latin America. Nature Communications. 2025;16: 8196. doi:10.1038/s41467-025-63277-6

28. Heeringa S, Connor J. Technical Description of the Health and Retirement Survey Sample Design. Ann Arbor, MI; 1995.

29. Faul J, Collins S, Smith J, Zhao W, Kardia S, Weir D. APOE and Serotonin Transporter Alleles – Early release. Ann Arbor, MI: University of Michigan; 2021. Available: https://hrsdata.isr.umich.edu/sites/default/files/documentation/data-descriptions/1629310147/File-Description-for-APOE-and-Serotonin.pdf

30. Sylvers DL, Hicken M, Esposito M, Manly J, Judd S, Clarke P. Walkable Neighborhoods and Cognition: Implications for the Design of Health Promoting Communities. Journal of Aging and Health. 2022;34: 893–904. doi:10.1177/08982643221075509

31. Brandt J, Spencer M, Folstein M. The telephone interview for cognitive status. Neuropsychiatry Neuropsychol Behav Neurol. 1988;1: 111–117.

32. Friedman EM, Shih RA, Slaughter ME, Weden MM, Cagney KA. Neighborhood age structure and cognitive function in a nationally-representative sample of older adults in the U.S. Social Science & Medicine. 2017;174: 149–158. doi:10.1016/j.socscimed.2016.12.005

33. Clarke P, Melendez R. National Neighborhood Data Archive (NaNDA): Neighborhood Socioeconomic and Demographic Characteristics of Census Tracts, United States, 2000-2010. ICPSR; 2019. 10.3886/E111107V1

34. Everitt BS, Landau S, Leese M, Stahl D. Cluster Analysis. Hoboken, NJ: John Wiley & Sons; 2011.

35. R Core Team. R: A language and environment for statistical computing. Vienna, Austria: R Foundation for Statistical Computing; 2021. Available: https://www.R-project.org/

36. StataCorp. Stata Statistical Software: Release 17. College Station, TX: StataCorp LLC.; 2021.

37. Powell LM, Slater S, Chaloupka FJ, Harper D. Availability of Physical Activity–Related Facilities and Neighborhood Demographic and Socioeconomic Characteristics: A National Study. Am J Public Health. 2006;96: 1676–1680. doi:10.2105/AJPH.2005.065573

38. Kim MH, Clarke P, Dunkle RE. Urban Neighborhood Characteristics and the Spatial Distribution of Home and Community-Based Service Organizations in Michigan Metropolitan Statistical Areas. Res Aging. 2022;44: 156–163. doi:10.1177/01640275211005079

39. Humphrey JL, Lindstrom M, Barton KE, Shrestha PM, Carlton EJ, Adgate JL, et al. Social and Environmental Neighborhood Typologies and Lung Function in a Low-Income, Urban Population. International Journal of Environmental Research and Public Health. 2019;16: 1133. doi:10.3390/ijerph16071133

40. Wong P-H, Kourtit K, Nijkamp P. The ideal neighbourhoods of successful ageing: A machine learning approach. Health & Place. 2021;72: 102704. doi:10.1016/j.healthplace.2021.102704

41. Scharlach AE. Aging in Context: Individual and Environmental Pathways to Aging-Friendly Communities—The 2015 Matthew A. Pollack Award Lecture. The Gerontologist. 2017;57: 606–618. doi:10.1093/geront/gnx017

42. Kelly CM, Schootman M, Baker EA, Barnidge EK, Lemes A. The association of sidewalk walkability and physical disorder with area-level race and poverty. Journal of Epidemiology and Community Health. 2007;61: 978 LP – 983. doi:10.1136/jech.2006.054775

43. Nguyen AW, Taylor HO, Lincoln KD, Qin W, Hamler T, Wang F, et al. Neighborhood Characteristics and Inflammation Among Older Black Americans: The Moderating Effects of Hopelessness and Pessimism. The Journals of Gerontology: Series A. 2022;77: 315–322. doi:10.1093/gerona/glab121

44. Qin W, Yu J, Brown T. Perceived Neighborhood Disorder, Health Behaviors, and Inflammation Among Older Adults: A Mediation Analysis. J Aging Health. 2025; 08982643251356732. doi:10.1177/08982643251356732

45. Howell NA, Farber S, Widener MJ, Allen J, Booth GL. Association between residential self-selection and non-residential built environment exposures. Health & Place. 2018;54: 149–154. doi:10.1016/j.healthplace.2018.08.009

46. Osypuk TL, Acevedo-Garcia D. Beyond individual neighborhoods: A geography of opportunity perspective for understanding racial/ethnic health disparities. Health & Place. 2010;16: 1113–1123. doi:10.1016/j.healthplace.2010.07.002

